# Outcome measures in implantable brain-computer interface research: a systematic review

**DOI:** 10.1101/2024.10.15.24315534

**Authors:** Esmee Dohle, Eleanor Swanson, Suraya Yusuf, Luka Jovanovic, Lucy Thompson, Hugo Layard Horsfall, William R Muirhead, Luke Bashford, Jamie Brannigan

**Author notes:** Corresponding author: Jamie Brannigan University of Oxford, Medical Sciences Division, Oxford OX3 9DU.

## Abstract

Implantable brain-computer interfaces (iBCIs) aim to restore function in patients with severe motor impairments by translating neural signals into motor outputs. As iBCI technology advances toward clinical application, assessing iBCI performance with robust and clinically relevant outcome measures becomes crucial. This systematic review analysed 77 studies, with 63.6% reporting outcome measures prospectively. Decoding outcomes were most frequently assessed (67.5%), followed by task performance (63.6%). Only 22.1% of studies reported a clinical outcome measure, often related to prosthetic limb function or activities of daily living. Successful iBCI translation and regulatory approval requires clinical outcomes developed collaboratively with individuals with motor impairments.

**One Sentence Summary:** Implantable brain-computer interface studies primarily evaluate engineering-related outcome measures over clinical outcome measures.

## INTRODUCTION

Brain-computer interfaces (BCIs) are systems which record neural signals and translate these into commands to control external devices, thereby bypassing dysfunctional or damaged pathways from brain to muscles. [1,2] The aim of motor BCIs is to restore functional independence for individuals with severe motor impairments, for instance due to amyotrophic lateral sclerosis (ALS), spinal cord injury (SCI), or stroke.

Following the first human microelectrode array implantation in 2004, [3] implantable BCIs (iBCIs) have enabled the control of computer cursors, digital clicks [4] and robotic prosthetic limbs. [5] More recently, implantable BCIs have been used to decode attempted handwriting [6] and attempted speech [7,8] in patients with paralysis. Alongside these advances in decoding and performance, fully implanted BCI systems have been developed, requiring substantially reduced setup burden and enabling independent use at home. [4,9–11] Despite advances in investigational clinical studies, no iBCI technology has yet received a full regulatory approval, or been adopted as a standard of care. A major challenge in the clinical translation of iBCI devices is an absence of consensus for clinically meaningful performance metrics that can be used when evaluating device efficacy in clinical trials. [12,13] In the USA, the Food and Drug Administration has highlighted the absence of an appropriate outcome measure, [14] and government funding has been awarded to investigate this. [15]

In this systematic review, we aim to assess the outcome measures reported in all iBCI publications. We aim to determine if any consensus can be identified from existing literature and to inform the future selection of iBCI clinical endpoints.

## RESULTS

### Search results

Our search identified a total of 4279 records across three databases (1652 in MEDLINE, 2197 in Embase and 430 in CINAHL). Following deduplication, 2711 records remained, of which 341 were selected for full-text screening. A total of 77 studies were included in the final analysis. A full Preferred Reporting Items for Systematic Reviews and Meta-Analysis (PRISMA) flowchart is shown in Figure 1.

**Figure 1:**
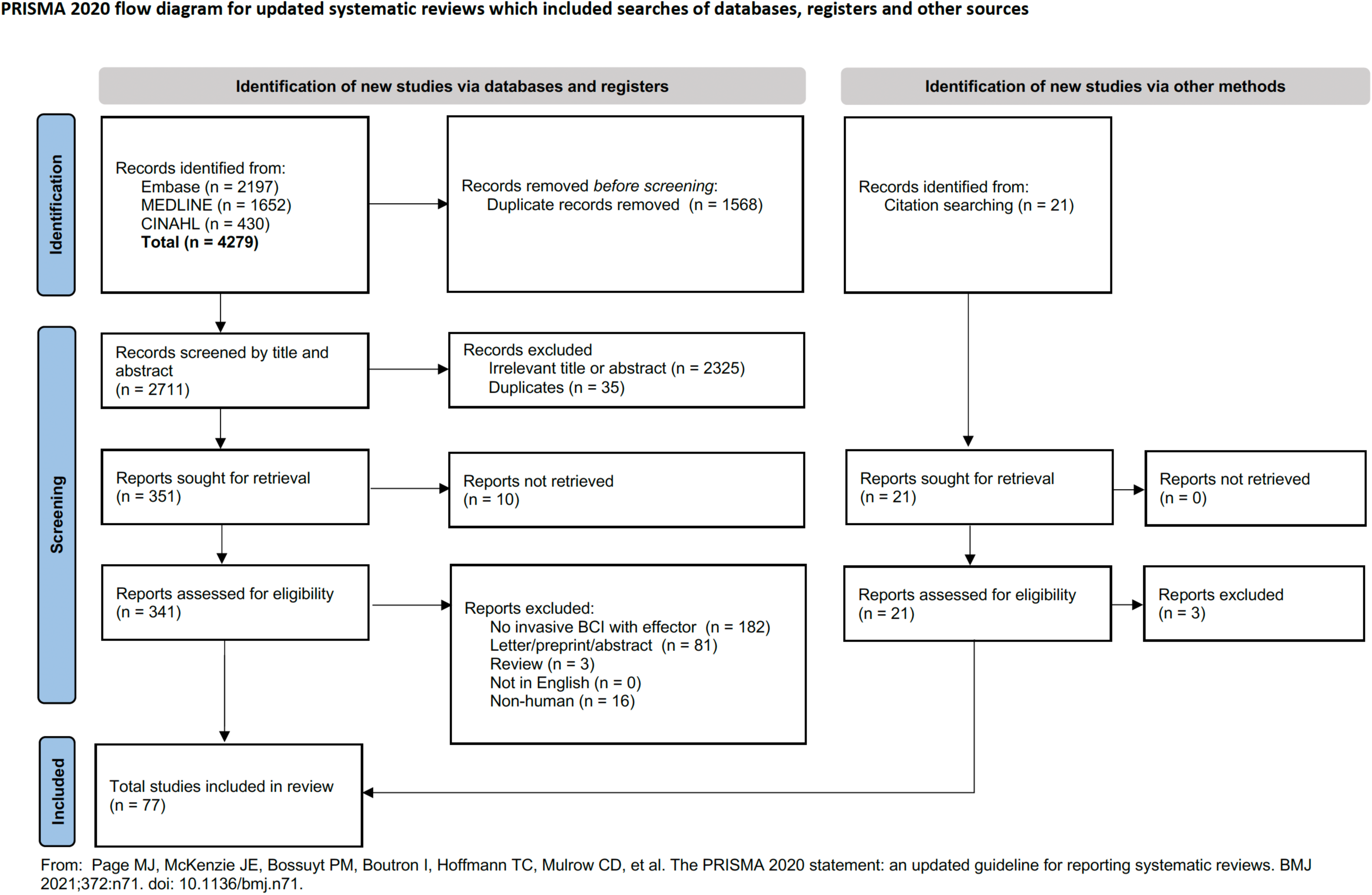
PRISMA flowchart.

### Study characteristics and participant demographics

The review included 77 studies published between 2000-2023. The majority (79%, n = 62) of studies were conducted in the United States. An overview of study characteristics is shown in Table 1. As some research participants were included in multiple publications, all publications were cross-referenced to identify 53 unique participants. The majority (77%, n = 41) of participants were male, with an average age of 46.2 years. A total of 45 patients (85%) suffered from amyotrophic lateral sclerosis (ALS), spinal cord injury (SCI) or stroke. An overview of participant demographics is shown in Table 2. Details of all included studies are shown in Supplementary Materials C. Most of the included studies, 62.8% (n = 49), reported their outcome measures prospectively.

**Table 1:** Overview of study characteristics.

|  |  |
| --- | --- |
| **Total** | **77** |
| Publication year (range) | 2000-2023 |
| Study design |  |
| Single-participant | 51 (66%) |
| Multi-participant | 26 (34%) |
| Country |  |
| USA | 62 |
| France | 4 |
| Netherlands | 3 |
| China | 2 |
| Switzerland | 2 |
| Australia | 2 |
| Canada | 2 |

**Table 2:** Overview of participant demographics.

|  |  |
| --- | --- |
| **Total participants** | **53** |
| Gender |  |
| Male | 41 (77%) |
| Female | 12 (23%) |
| Average age (earliest reported) | 46.2 |
| Pathology |  |
| Spinal cord injury (SCI) | 21 (40%) |
| Amyotrophic lateral sclerosis (ALS) | 16 (30%) |
| Stroke | 8 (15%) |
| Other | 8 (15%) |

### Types of outcome measures reported

Among the different categories of outcome measures, decoding-related outcomes such as decoding accuracy were the most frequently reported, in 67.5% of the publications (52 publications). Task-related outcomes, such as successful task completion or target accuracy, were also reported most publications (63.6%, n = 49). Clinical outcomes were more rarely used, with only 22.1% (17 publications) reporting a clinical outcome. This is shown in Figure 2.

**Figure 2:**
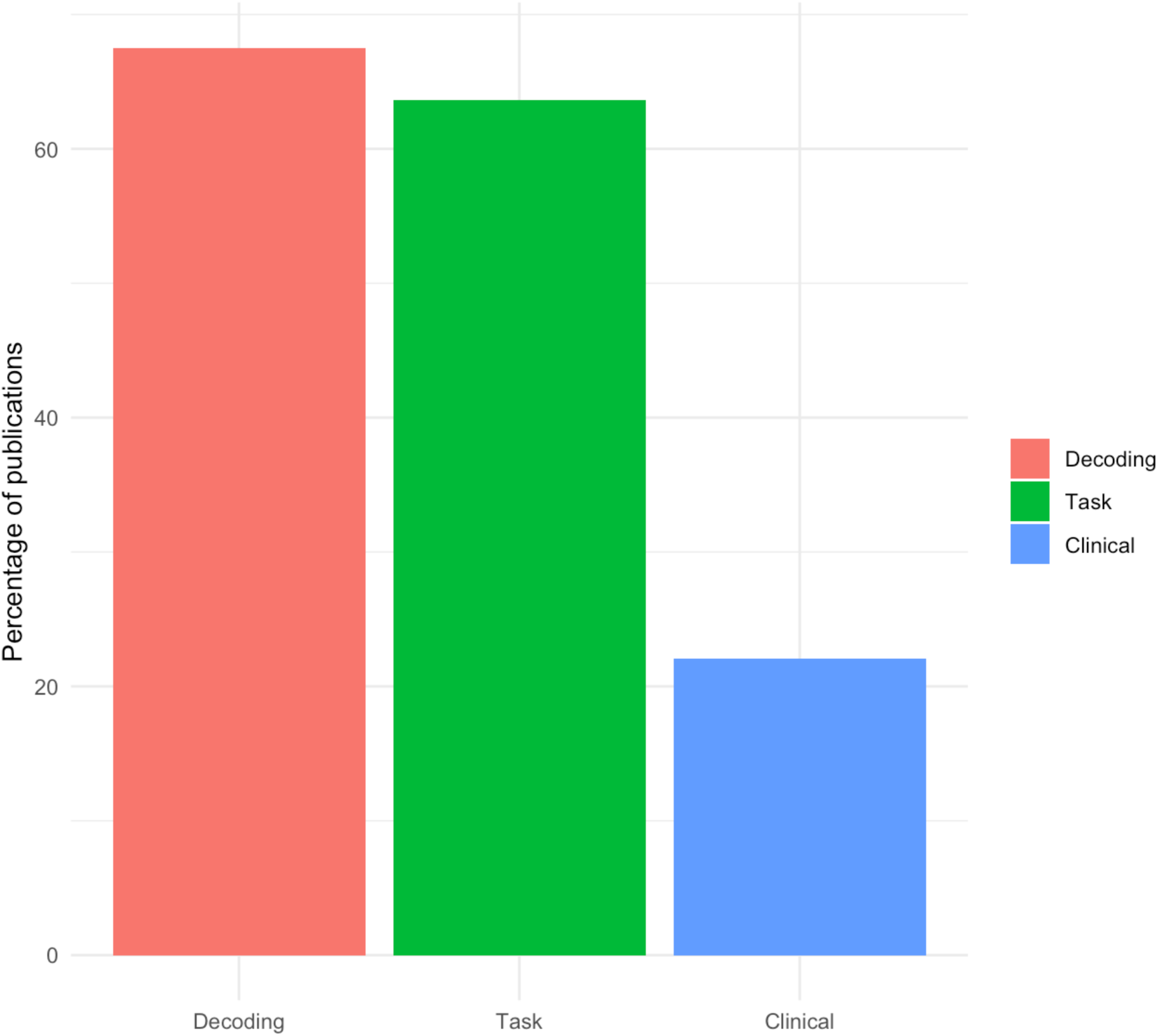
Percentage of publications reporting decoding-related, task-related and clinical outcome measures.

### Clinical outcome measures

The clinical outcome measures used varied widely, with 20 different clinical outcomes reported across 17 publications. Nearly half (47%, n = 8) of the studies reporting a clinical outcome were published after 2020.

Clinical outcome measures most commonly related to upper limb functioning, such as the Action Research Arm Test (ARAT) or the Graded and Redefined Assessment of Strength, Sensibility and Prehension (GRASSP), used by 13 publications. The second most commonly assessed type of clinical outcome were activities of daily living (ADLs), such as communication, online banking and shopping, with 6 publications assessing completion of at least one ADL. Other outcome measures relate to assistive device functioning, such as the Psychosocial Impact of Assistive Devices (PIADS) scale, adverse events and quality of life (QOL). This is shown in Figure 3.

**Figure 3:**
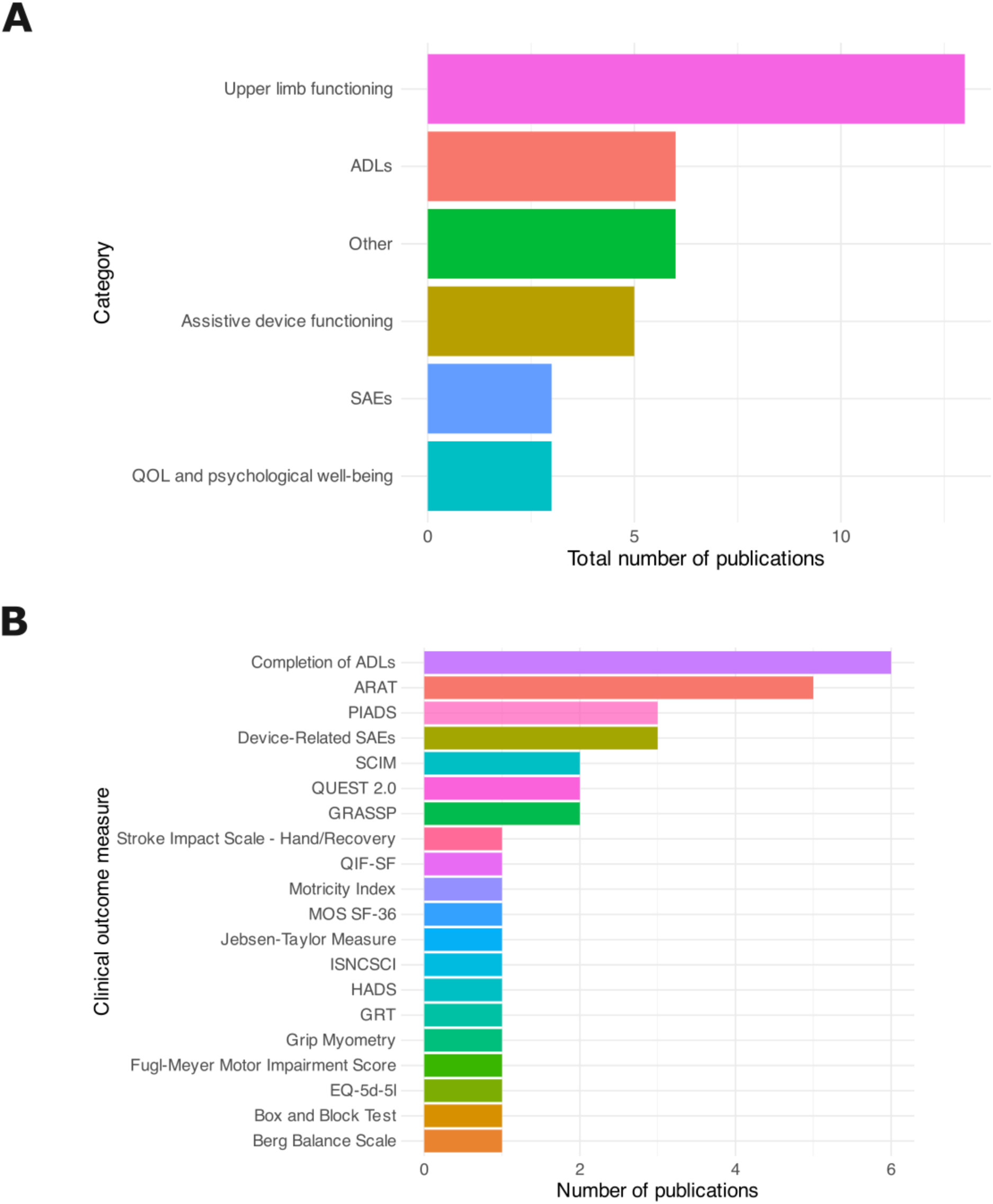
(A) Number of publications reporting different clinical outcome measures, grouped by category. (B) Number of publications reporting different individual clinical outcome measures. ARAT = Action Research Arm Test. BBS = Berg Balance Scale. BBT = Box and Block Test. EQ-5d-5l = EuroQOL-5d-5l. FMMIS = Fugl-Meyer Motor Impairment Score. GRASSP = Graded and Redefined Assessment of Strength Sensibility and Prehension. GRT = Grasp Release Test. HADS = Hospital Anxiety and Depression Scale. ISNCSCI = International Standards for Neurological Classification of Spinal Cord Injury. MOS SF-36 = Medical Outcomes Study Short Form-36. PIADS = Psychosocial Impact of Assistive Devices Scales. QIF-SF = Quadriplegia Index of Function - Short Form. QUEST 2.0 = Quebec User Evaluation of Satisfaction with Assistive Technology version 2.0. SCIM = Spinal Cord Independence Measure.

### Decoding- and task-related outcome measures

The most commonly reported engineering outcome measure was accuracy, with 76% of of the included studies (59 publications) reporting this as an outcome. Task accuracy was most commonly reported, e.g. task success rate or accuracy (47%, n = 36) followed by model accuracy, e.g. decoding or classification accuracy (44%, n = 34).

Several studies involved iBCIs developed for the purpose of assisting communication, e.g. via cursor-based typing or speech/phoneme decoding. Out of 78 total studies, 14 studies reported a communication-speed outcome measure such as correct characters per minute (CCPM) or words per minute (WPM). Of these studies, the majority (8 publications) used character-based metrics such as CCPM or CPM, although more recent studies (6 publications) tend to use word-based metrics such as WPM.

Another metric of speed, the information transfer rate (ITR) and its derivatives, was reported in 8 studies (10% of all publications). Combined, a total of 18 studies (23%) reported a speed-related metric such as CPM or ITR.

## DISCUSSION

To our knowledge, this is the first systematic review of outcome measures used to assess iBCI devices. We identified a total population of 53 participants with iBCIs evaluated across 77 studies. Most studies assessed iBCI outcomes using measures of engineering performance, such as decoder or task performance. Only 22.1% (n = 17) studies reported a clinical outcome measure, most of which evaluated robotic prosthetic upper limb function (n = 13). The proportion of studies evaluating clinical iBCI outcomes has increased in recent years, however clinical measures were heterogenous and often specific to the types of tasks being performed.

### Clinical iBCI outcome measures are increasingly utilised, but highly heterogenous

Of the 18 studies we identified utilising clinical outcome measures, nearly half (47%, n = 8) were published since 2020, and this represents a higher proportion of the outcome assessments being used to assess iBCI devices. Despite increasing interest in evaluating the clinical benefit of iBCI devices, there is substantial variability in the assessments being used. This has included quality of life (QoL) metrics, specific measures of upper limb function, and assessments of satisfaction in the use of assistive devices.

In most cases, assessments were specific to a single task or function being restored, most notably in the case of restored upper limb function. Whilst these measures may be useful in a specific context, such measures are neither agnostic to task nor device, precluding use as a generalised measure of iBCI outcomes. Moreover, assessments of physical function are less immediately relevant, as ongoing iBCI studies are primarily aiming to restore control of digital devices and/or communication (e.g. BrainGate2 (NCT00912041), Neuralink PRIME (NCT06429735), Synchron COMMAND (NCT05035823), BRAVO (NCT03698149)). This suggests outcome measures which can capture the clinical benefits of restored digital functional independence are most appropriate for the first wave of iBCI devices approaching clinical translation.

The single most commonly captured clinical outcome was the performance of activities of daily living. However, this typically comprised assessment of individual activities rather than using an existing standardised measure. To our knowledge, no ADL currently exists which captures restored digital functional independence. The development of a ‘digital activities of daily living’ instrument has previously been proposed by Fry et al., and the US Food and Drug Administration. [12,13].

The quality of life (QoL) measures identified in our review, such as the EuroQol-5D-5L, are existing measures which are agnostic to both device type and the function being restored. QoL assessments measure an individual’s perception of their overall well-being, which includes feelings about health status and the nonmedical aspects of one’s life. However, QoL assessments are typically used only as supplementary measures when evaluating therapeutic interventions due to inherent limitations. They are confounded by a wide range of variables, such as socioeconomic status and psychological wellbeing, leading to temporal fluctuations and a high degree of inter-subject variability. If constricted to health-related QoL, this is still confounded by comorbidities. Moreover, a study of patient perceptions in ALS, a population involved in current BCI studies, has demonstrated persistently elevated QoL, despite progressive paralysis of all four limbs and the resulting dependence upon carers. [16] This suggests an effective psychological adaptation to new deficits, which would skew the assessment of any intervention to restore bodily functions. Whilst it is necessary to capture patient reported outcomes in randomised studies of interventions, [17] and a QoL measure may be the most such appropriate assessment, these challenges may limit the potential for QoL assessments as a primary measure of clinical benefit with iBCI devices.

Given the inherent heterogeneity in device types, desired outputs, patient selection, and baseline motor impairments, it is unlikely that one single comprehensive measure will be developed to evaluate overall iBCI clinical benefit. To this end, Fry *et al.* (2022) suggest that BCI clinical outcome measures should address three dimensions: how a patient ‘feels’ (e.g. quality of life scores), how a patient ‘functions’ (e.g. activities of daily living scale) and how a patient ‘survives’ (i.e. health-related outcomes, including device safety). [12]

### Engineering performance measures have been selected with greater consistency

Engineering measures of decoder and task performance were more commonly and consistently used in the included studies. Accuracy was measured in 76% of studies (n=59). Performance measures related to speed, e.g. characters per minute or bit rate, were utilised in only 23% of studies (n = 18). The preference for measuring device accuracy as an engineering metric is aligned with patient preference literature, which demonstrates patients prioritise high accuracy over other aspects of iBCI performance, such as speed. Whilst commonly used and preferred by patients as a performance characteristic, accuracy is a one dimensional measure and does not give information on the task difficulty or complexity, e.g. the degrees of freedom, cognitive load being used, or training burden. Moreover, isolated measurement of accuracy does not account for the environmental context in which a task can be performed. Therefore, such engineering measures do not account for how an individual feels and functions in their daily life, [18] and the US Food and Drug Administration have referred to these assessments as ‘lab-tests’, rather than assessments of real world function. [13] Inclusion of engineering metrics, such as accuracy, may still be useful to evaluate performance of the BCI device, however additional assessments of real world function will be necessary to determine clinical beneifts.

Accuracy was most commonly defined at the task level, i.e., number of successful trials/total number of trials, with assessment in 36 studies (47%). This definition at the task level is most appropriate for application across different iBCI studies, as it is a standardised accuracy measure for discrete, continuous, and hybrid discrete/continuous iBCI applications. Other definitions, such as classification accuracy, are specific to the nature of the paradigm employed when decoding motor intentions.

Whilst information transfer rate or characters per minute of language output have also been referenced as measures of BCI function, [13] these assessments were overrepresented by benchmark studies of iBCI performance, and less commonly used in the literature overall. However, it may be the case that these measures are only reported if successful in demonstrating a new breakthrough, as there is less frequent reporting on low or failed outcome measures across the academic literature.

### Towards a standardised and clinically meaningful iBCI outcome measures

Current iBCI research primarily utilises engineering-related outcome measures, quantifying device performance in terms of decoding accuracy or subsequent performance at tasks. This focus is understandable, given the considerable ongoing and addressed technical challenges in developing iBCI systems. Furthermore, device-related outcomes will remain important in the context of fundamental science research, iBCI quality control, and development of next generation devices. However, to develop and validate devices for clinical translation, an increased focus on clinically meaningdul, patient-centred concepts of interest is essential. [13,18] This may include evaluations of functional independence, patient quality of life, or direct assessments of restored bodily functions.

The selection and development of appropriate clinical outcome assessments must involve multi-stakeholder consensus, [13] including input from individuals with lived experience of severe motor impairment. This is critical to ensure future device outcomes match a user’s priorities and expectations. Moreover, given the emergence of multiple companies working to translate iBCI devices at scale, this work must also ensure collaboration across different commercial actors. In the USA, the creation of the iBCI collaborative community (iBCI-CC) is an important step to enable this work (ibci-cc.org), along with organisation of multiple workshops by regulators to discuss iBCI clinical outcome assessments. [13,14]

## MATERIALS AND METHODS

This systematic review was conducted in line with Preferred Reporting Items for Systematic Reviews and Meta-Analysis (PRISMA) 2020 guidelines. A PRISMA checklist is attached in Supplementary Materials A. The review was prospectively registered on Open Science Framework (OSF) (https://osf.io/ky5g4).

### Search strategy

Search strategies were developed for three databases (MEDLINE, Embase and CINAHL) which combined synonyms for brain-computer interfaces, intracortical and patient. A senior medical librarian was consulted throughout the process. The search was carried out using Ovid (Wolters Kluwer, Netherlands) and EBSCO, and run from inception to 12^th^ December 2023. An example search strategy is included in Supplementary Materials B. Further studies were identified through reference lists of included records.

### Eligibility criteria

Screening was performed in accordance with the criteria shown in Table 3. The scope of this review was limited to implantable BCI devices, defined as an intracranial device which records neural activity and decodes this into an output signal to control an external effector. Non-invasive BCI devices and cochlear implants have previously been discussed in detail. [19–23] Additionally, studies were excluded where transient implantation was carried out for the primary purpose of peri-operative care, e.g. seizure localization in epilepsy patients.

**Table 3:**
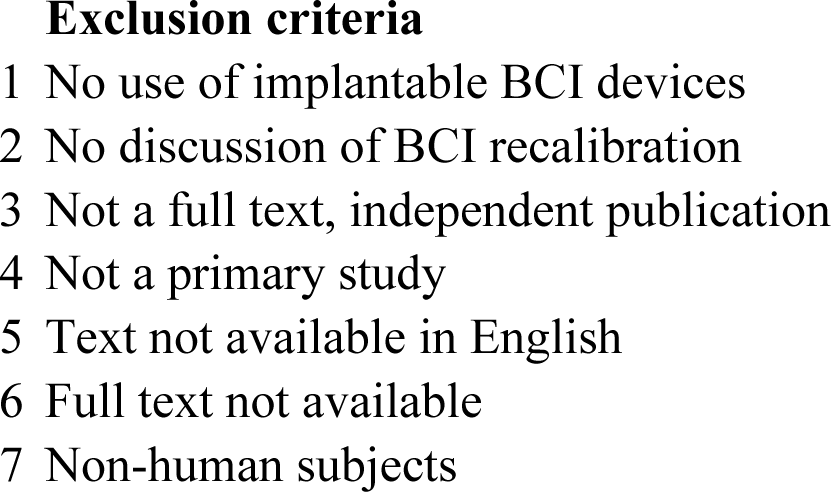
Exclusion criteria.

### Study selection

Included records were screened by two independent reviewers (ED, ES). An initial pilot screen of 50 records was carried out to ensure concordance, following which reviewers were blinded to each other’s decisions using Rayyan (Rayyan Systems Inc, Cambridge, MA, USA). Disagreements were resolved by consensus or discussion with a third reviewer (JB). Full-text screening was carried out by the same reviewers (ED, ES).

### Data management, extraction, and appraisal

Data extraction was carried out using a piloted proforma in Microsoft Excel (Microsoft, Redmond, WA, USA). Risk of bias assessment was carried out in duplicate by two reviewers (ES, ED) using the Mixed Methods Appraisal Tool (MMAT) checklist. Zotero (Zotero, Vienna, VA, USA) was used for reference management.

### Data synthesis

Analysis was carried out in R (R Core Team, 2019). Plots were produced using the ggplot2 package. [24] Figures were edited using Inkscape (Inkscape Project, 2020).

As protocol papers were rarely published prior to publication of the included manuscripts, outcome measures were considered to be prospectively reported if they were clearly defined in the methods section.

During data extraction, outcome measures were divided into three categories: 1. evaluation of decoder performance (e.g. classifier accuracy), 2. evaluation of task performance (e.g. successful target acquisition), or 3. assessment of clinical outcome (e.g. Graded Redefined Assessment of Strength, Sensibility and Prehension, GRASSP scale).

## Data Availability

Data are available on reasonable request.

## List of Supplementary Materials

Supplementary Materials A: PRISMA checklist

Supplementary Materials B: Example search strategy as applied to MEDLINE

Supplementary Materials C: Table of included studies

## Acknowledgments

N/A

## Author contributions

Conceptualization: JB, LB, ED, ES

Methodology: ED, ES, JB, LB

Visualization: ED, JB, LJ

Data extraction: ED, ES, SY, LJ, LT

Writing – original draft: ED, ES, HLH, LB, JB

Writing – review & editing: ED, ES, HLH, LB, JB, LJ, SY, LT

## Data and materials availability

Data are available on reasonable request.

