## Supplementary Materials for "Outcome measures in implantable brain-computer interface research: a systematic review"

**List of Supplementary Materials**

Supplementary Materials A: PRISMA checklist
Supplementary Materials B: Example search strategy as applied to MEDLINE
Supplementary Materials C: Table of included studies

**Supplementary materials A: PRISMA checklist**

| Section and Topic | Item # | Checklist item | Location where item is reported |
|---|---|---|---|
| **TITLE** | | | |
| Title | 1 | Identify the report as a systematic review. | 1 |
| **ABSTRACT** | | | |
| Abstract | 2 | See the PRISMA 2020 for Abstracts checklist. | 2 |
| **INTRODUCTION** | | | |
| Rationale | 3 | Describe the rationale for the review in the context of existing knowledge. | 3 |
| Objectives | 4 | Provide an explicit statement of the objective(s) or question(s) the review addresses. | 3 |
| **METHODS** | | | |
| Eligibility criteria | 5 | Specify the inclusion and exclusion criteria for the review and how studies were grouped for the syntheses. | 17-19 |
| Information sources | 6 | Specify all databases, registers, websites, organisations, reference lists and other sources searched or consulted to identify studies. Specify the date when each source was last searched or consulted. | 17-19 |
| Search strategy | 7 | Present the full search strategies for all databases, registers and websites, including any filters and limits used. | Supplementary Materials B |
| Selection process | 8 | Specify the methods used to decide whether a study met the inclusion criteria of the review, including how many reviewers screened each record and each report retrieved, whether they worked independently, and if applicable, details of automation tools used in the process. | 17-19 |
| Data collection process | 9 | Specify the methods used to collect data from reports, including how many reviewers collected data from each report, whether they worked independently, any processes for obtaining or confirming data from study investigators, and if applicable, details of automation tools used in the process. | 17-19 |
| Data items | 10a | List and define all outcomes for which data were sought. Specify whether all results that were compatible with each outcome domain in each study were sought (e.g. for all measures, time points, analyses), and if not, the methods used to decide which results to collect. | 17-19 |
| | 10b | List and define all other variables for which data were sought (e.g. participant and intervention characteristics, funding sources). Describe any assumptions made about any missing or unclear information. | |
| Study risk of bias assessment | 11 | Specify the methods used to assess risk of bias in the included studies, including details of the tool(s) used, how many reviewers assessed each study and whether they worked independently, and if applicable, details of automation tools used in the process. | 17-19 |
| Effect measures | 12 | Specify for each outcome the effect measure(s) (e.g. risk ratio, mean difference) used in the synthesis or presentation of results. | N/A |
| Synthesis methods | 13a | Describe the processes used to decide which studies were eligible for each synthesis (e.g. tabulating the study intervention characteristics and comparing against the planned groups for each synthesis (item #5)). | 17-19 |
| | 13b | Describe any methods required to prepare the data for presentation or synthesis, such as handling of missing summary statistics, or data conversions. | 17-19 |

| Section and Topic | Item # | Checklist item | Location where item is reported |
|---|---|---|---|
| | 13c | Describe any methods used to tabulate or visually display results of individual studies and syntheses. | 17-19 |
| | 13d | Describe any methods used to synthesize results and provide a rationale for the choice(s). If meta-analysis was performed, describe the model(s), method(s) to identify the presence and extent of statistical heterogeneity, and software package(s) used. | 17-19 |
| | 13e | Describe any methods used to explore possible causes of heterogeneity among study results (e.g. subgroup analysis, meta-regression). | 17-19 |
| | 13f | Describe any sensitivity analyses conducted to assess robustness of the synthesized results. | N/A |
| Reporting bias assessment | 14 | Describe any methods used to assess risk of bias due to missing results in a synthesis (arising from reporting biases). | |
| Certainty assessment | 15 | Describe any methods used to assess certainty (or confidence) in the body of evidence for an outcome. | |
| **RESULTS** | | | |
| Study selection | 16a | Describe the results of the search and selection process, from the number of records identified in the search to the number of studies included in the review, ideally using a flow diagram. | 4-5 |
| | 16b | Cite studies that might appear to meet the inclusion criteria, but which were excluded, and explain why they were excluded. | 5, Methods |
| Study characteristics | 17 | Cite each included study and present its characteristics. | Supplementary Materials C |
| Risk of bias in studies | 18 | Present assessments of risk of bias for each included study. | MMAT results on request |
| Results of individual studies | 19 | For all outcomes, present, for each study: (a) summary statistics for each group (where appropriate) and (b) an effect estimate and its precision (e.g. confidence/credible interval), ideally using structured tables or plots. | 4-11 |
| Results of syntheses | 20a | For each synthesis, briefly summarise the characteristics and risk of bias among contributing studies. | |
| | 20b | Present results of all statistical syntheses conducted. If meta-analysis was done, present for each the summary estimate and its precision (e.g. confidence/credible interval) and measures of statistical heterogeneity. If comparing groups, describe the direction of the effect. | 8-15 |
| | 20c | Present results of all investigations of possible causes of heterogeneity among study results. | MMAT results on request |
| | 20d | Present results of all sensitivity analyses conducted to assess the robustness of the synthesized results. | |
| Reporting biases | 21 | Present assessments of risk of bias due to missing results (arising from reporting biases) for each synthesis assessed. | MMAT results on request |
| Certainty of evidence | 22 | Present assessments of certainty (or confidence) in the body of evidence for each outcome assessed. | MMAT results on request |
| **DISCUSSION** | | | |
| Discussion | 23a | Provide a general interpretation of the results in the context of other evidence. | 12-16 |

| Section and Topic | Item # | Checklist item | Location where item is reported |
|---|---|---|---|
| | 23b | Discuss any limitations of the evidence included in the review. | 12-16 |
| | 23c | Discuss any limitations of the review processes used. | 12-16 |
| | 23d | Discuss implications of the results for practice, policy, and future research. | 12-16 |
| **OTHER INFORMATION** | | | |
| Registration and protocol | 24a | Provide registration information for the review, including register name and registration number, or state that the review was not registered. | 17 |
| | 24b | Indicate where the review protocol can be accessed, or state that a protocol was not prepared. | 17 |
| | 24c | Describe and explain any amendments to information provided at registration or in the protocol. | N/A |
| Support | 25 | Describe sources of financial or non-financial support for the review, and the role of the funders or sponsors in the review. | 31 |
| Competing interests | 26 | Declare any competing interests of review authors. | 31 |
| Availability of data, code and other materials | 27 | Report which of the following are publicly available and where they can be found: template data collection forms; data extracted from included studies; data used for all analyses; analytic code; any other materials used in the review. | Available on request |

*From:* Page MJ, McKenzie JE, Bossuyt PM, Boutron I, Hoffmann TC, Mulrow CD, et al. The PRISMA 2020 statement: an updated guideline for reporting systematic reviews. BMJ 2021;372:n71. doi: 10.1136/bmj.n71

**Supplementary materials B: Example search strategy as applied to MEDLINE**

| | | |
|---|---|---|
| 1 | Brain-Computer Interfaces/ | 4689 |
| 2 | brain computer interface*.mp. | 8312 |
| 3 | BCI*.mp. | 7982 |
| 4 | brain machine interface*.mp. | 1790 |
| 5 | neural prosthe*.mp. | 1346 |
| 6 | neuroprosthe*.mp. | 2075 |
| 7 | neuromotor prosthe*.mp. | 19 |
| 8 | 1 or 2 or 3 or 4 or 5 or 6 or 7 | 14387 |
| 9 | implant*.mp. | 608877 |
| 10 | "prostheses and implants"/ | 50025 |
| 11 | electrodes, implanted/ | 21495 |
| 12 | neural prostheses/ | 503 |
| 13 | intracortical.mp. | 7143 |
| 14 | microelectrode array*.mp. | 2730 |
| 15 | MEA.mp. | 4324 |
| 16 | (Utah adj2 array).mp. | 72 |
| 17 | electrocorticography.mp. | 3409 |
| 18 | ecog.mp. | 10740 |
| 19 | stentrode*.mp. | 19 |
| 20 | endovascular.mp. | 75471 |
| 21 | 9 or 10 or 11 or 12 or 13 or 14 or 15 or 16 or 17 or 18 or 19 or 20 | 689516 |
| 22 | patient*.mp. | 8670314 |
| 23 | user*.mp. | 299593 |
| 24 | individual*.mp. | 2031510 |
| 25 | exp Patients/ | 84088 |
| 26 | 22 or 23 or 24 or 25 | 10195016 |
| 27 | 8 and 21 and 26 | 1652 |

**Supplementary materials C: Table of included studies**

| Year | Authors | Study title |
|---|---|---|
| 2015 | Aflalo et al. [25] | Neurophysiology. Decoding motor imagery from the posterior parietal cortex of a tetraplegic human. |
| 2017 | Ajiboye et al. [26] | Restoration of reaching and grasping movements through brain-controlled muscle stimulation in a person with tetraplegia: a proof-of-concept demonstration. |
| 2015 | Bacher et al. [27] | Neural Point-and-Click Communication by a Person With Incomplete Locked-In Syndrome. |
| 2019 | Benabid et al. [28] | An exoskeleton controlled by an epidural wireless brain-machine interface in a tetraplegic patient: a proof-of-concept demonstration. |
| 2019 | Bockbrader et al. [29] | Clinically Significant Gains in Skillful Grasp Coordination by an Individual With Tetraplegia Using an Implanted Brain-Computer Interface With Forearm Transcutaneous Muscle Stimulation. |
| 2016 | Bouton et al. [30] | Restoring cortical control of functional movement in a human with quadriplegia |
| 2018 | Brandman et al. [31] | Robust Closed-Loop Control of a Cursor in a Person with Tetraplegia using Gaussian Process Regression. |
| 2018 | Brandman et al.[32] | Rapid calibration of an intracortical brain-computer interface for people with tetraplegia. |
| 2021 | Cajigas et al. [33] | Implantable brain–computer interface for neuroprosthetic-enabled volitional hand grasp restoration in spinal cord injury |
| 2023 | Cajigas et al. [34] | Brain-Computer interface control of stepping from invasive electrocorticography upper-limb motor imagery in a patient with quadriplegia. |
| 2011 | Chadwick et al. [35] | Continuous neuronal ensemble control of simulated arm reaching by a human with tetraplegia |
| 2013 | Collinger et al. [36] | High-performance neuroprosthetic control by an individual with tetraplegia. |
| 2022 | Davis et al. [37] | Design-development of an at-home modular brain-computer interface (BCI) platform in a case study of cervical spinal cord injury. |
| 2018 | Degenhart et al. [38] | Remapping cortical modulation for brain-machine interfaces: A somatotopy-based approach in individuals with upper-limb paralysis |
| 2021 | Dekleva et al. [39] | Generalizable cursor click decoding using grasp-related neural transients. |
| 2016 | Downey et al. [40] | Blending of brain-machine interface and vision-guided autonomous robotics improves neuroprosthetic arm performance during grasping |
| 2018 | Downey et al. [41] | Implicit Grasp Force Representation in Human Motor Cortical Recordings. |
| 2022 | Feng et al. [42] | Design a Novel BCI for Neurorehabilitation Using Concurrent LFP and EEG Features: A Case Study. |
| 2017 | Friedenberg et al. [43] | Neuroprosthetic-enabled control of graded arm muscle contraction in a paralyzed human. |

| | | |
|---|---|---|
| 2015 | Gilja et al. [44] | Clinical translation of a high-performance neural prosthesis. |
| 2023 | Guan et al. [45] | Decoding and geometry of ten finger movements in human posterior parietal cortex and motor cortex. |
| 2022 | Guan et al. [46] | Stability of motor representations after paralysis. |
| 2009 | Guenther et al. [47] | A wireless brain-machine interface for real-time speech synthesis. |
| 2022 | Guthrie et al. [48] | The impact of distractions on intracortical brain–computer interface control of a robotic arm |
| 2022 | Handelman et al. [49] | Shared Control of Bimanual Robotic Limbs With a Brain-Machine Interface for Self-Feeding. |
| 2006 | Hochberg et al. [3] | Neuronal ensemble control of prosthetic devices by a human with tetraplegia. |
| 2012 | Hochberg et al. [50] | Reach and grasp by people with tetraplegia using a neurally controlled robotic arm. |
| 2015 | Jarosiewicz et al. [51] | Virtual typing by people with tetraplegia using a self-calibrating intracortical brain-computer interface. |
| 2016 | Jarosiewicz et al. [52] | Retrospectively supervised click decoder calibration for self-calibrating point-and-click brain-computer interfaces. |
| 2022 | Jiang et al. [53] | Short report: surgery for implantable brain-computer interface assisted by robotic navigation system. |
| 2020 | Jorge et al. [54] | Classification of Individual Finger Movements Using Intracortical Recordings in Human Motor Cortex. |
| 2004 | Kennedy et al. [55] | Computer control using human intracortical local field potentials |
| 2000 | Kennedy et al. [56] | Direct control of a computer from the human central nervous system. |
| 2011 | Kennedy et al. [57] | Making the lifetime connection between brain and machine for restoring and enhancing function. |
| 2011 | Kim et al. [58] | Point-and-click cursor control with an intracortical neural interface system by humans with tetraplegia. |
| 2008 | Kim et al. [59] | Neural control of computer cursor velocity by decoding motor cortical spiking activity in humans with tetraplegia |
| 2017 | Kryger et al. [60] | Flight simulation using a Brain-Computer Interface: A pilot, pilot study. |
| 2020 | Leinders et al. [61] | Dorsolateral prefrontal cortex-based control with an implanted brain-computer interface. |
| 2023 | Lorach et al. [62] | Walking naturally after spinal cord injury using a brain-spine interface. |
| 2023 | Luo et al. [63] | Stable Decoding from a Speech BCI Enables Control for an Individual with ALS without Recalibration for 3 Months. |
| 2012 | Marquez-Chin et al. [64] | Real-time two-dimensional asynchronous control of a computer cursor with a single subdural electrode. |
| 2014 | Masse et al. [65] | Non-causal spike filtering improves decoding of movement intention for intracortical BCIs |
| 2022 | Metzger et al. [66] | Generalizable spelling using a speech neuroprosthesis in an individual with severe limb and vocal paralysis |
| 2023 | Metzger et al. [8] | A high-performance neuroprosthesis for speech decoding and avatar control |

| | | |
|---|---|---|
| 2023 | Mitchell et al. [4] | Assessment of Safety of a Fully Implanted Endovascular Brain-Computer Interface for Severe Paralysis in 4 Patients: The Stentrode With Thought-Controlled Digital Switch (SWITCH) Study. |
| 2022 | Moly et al. [67] | An adaptive closed-loop ECoG decoder for long-term and stable bimanual control of an exoskeleton by a tetraplegic. |
| 2021 | Moses et al. [7] | Neuroprosthesis for Decoding Speech in a Paralyzed Person with Anarthria |
| 2018 | Nuyujukian et al. [68] | Cortical control of a tablet computer by people with paralysis |
| 2021 | Oxley et al. [69] | Motor neuroprosthesis implanted with neurointerventional surgery improves capacity for activities of daily living tasks in severe paralysis: first in-human experience |
| 2017 | Pandarinath et al. [70] | High performance communication by people with paralysis using an intracortical brain-computer interface |
| 2019 | Pels et al. [71] | Stability of a chronic implanted brain-computer interface in late-stage amyotrophic lateral sclerosis. |
| 2013 | Perge et al. [72] | Intra-day signal instabilities affect decoding performance in an intracortical neural interface system. |
| 2014 | Perge et al. [73] | Reliability of directional information in unsorted spikes and local field potentials recorded in human motor cortex. |
| 2023 | Rizzoglio et al. [74] | From monkeys to humans: observation-basedEMGbrain-computer interface decoders for humans with paralysis. |
| 2022 | Rouanne et al. [75] | Unsupervised adaptation of an ECoG based brain-computer interface using neural correlates of task performance. |
| 2023 | Rubin et al. [76] | Interim Safety Profile From the Feasibility Study of the BrainGate Neural Interface System. |
| 2022 | Rubin et al. [77] | Learned Motor Patterns Are Replayed in Human Motor Cortex during Sleep |
| 2019 | Sakellaridi et al. [78] | Intrinsic Variable Learning for Brain-Machine Interface Control by Human Anterior Intraparietal Cortex. |
| 2022 | Serino et al. [79] | Sense of agency for intracortical brain-machine interfaces. |
| 2022 | Serruya et al. [80] | Neuromotor prosthetic to treat stroke-related paresis: N-of-1 trial. |
| 2023 | Shah et al. [81] | A brain-computer typing interface using finger movements. |
| 2013 | Shaikhouni et al. [82] | Somatosensory responses in a human motor cortex |
| 2021 | Silversmith et al. [83] | Plug-and-play control of a brain–computer interface through neural map stabilization |
| 2011 | Simeral et al. [84] | Neural control of cursor trajectory and click by a human with tetraplegia 1000 days after implant of an intracortical microelectrode array. |
| 2021 | Simeral et al. [85] | Home Use of a Percutaneous Wireless Intracortical Brain-Computer Interface by Individuals With Tetraplegia. |
| 2022 | Sliwowski et al. [86] | Decoding ECoG signal into 3D hand translation using deep learning. |

| | | |
|---|---|---|
| 2017 | Talakoub et al. [87] | Reconstruction of upper limb movement from electrocorticographic signals to control functional electrical stimulation for hand function restoration |
| 2008 | Truccolo et al. [88] | Primary Motor Cortex Tuning to Intended Movement Kinematics in Humans with Tetraplegia |
| 2016 | Vansteensel et al. [89] | Fully Implanted Brain-Computer Interface in a Locked-In Patient with ALS. |
| 2013 | Wang et al. [5] | An electrocorticographic brain interface in an individual with tetraplegia. |
| 2019 | Weiss et al. [90] | Demonstration of a portable intracortical brain-computer interface |
| 2019 | Willett et al. [91] | Principled BCI Decoder Design and Parameter Selection Using a Feedback Control Model. |
| 2021 | Willett et al. [6] | High-performance brain-to-text communication via handwriting |
| 2023 | Willett et al. [92] | A high-performance speech neuroprosthesis |
| 2014 | Wodlinger et al. [93] | Ten-dimensional anthropomorphic arm control in a human brain-machine interface: difficulties, solutions, and limitations |
| 2018 | Young et al. [94] | Signal processing methods for reducing artifacts in microelectrode brain recordings caused by functional electrical stimulation |
| 2020 | Zhang et al. [95] | Preservation of Partially Mixed Selectivity in Human Posterior Parietal Cortex across Changes in Task Context. |
